# External validation of the Meggitt-Wagner, Texas University, SINBAD, and Saint Elian classifications for predicting major amputation in patients with diabetes at a public hospital in Peru

**DOI:** 10.1101/2025.06.19.25329964

**Authors:** Luis Alberto Gallardo-Alburqueque, Yudith Quispe-Landeo, Ann Chanamé-Marín, Leonardo J. Uribe-Cavero, Marlon Yovera-Aldana

## Abstract

**Objective:** To compare the prognostic discriminative capacity of the Meggitt-Wagner, University of Texas, SINBAD, and Saint Elian classifications for predicting major amputation in patients with diabetes treated at a public hospital in Peru.

**Materials and Methods:** A retrospective cohort study was conducted, including patients with a follow-up period of up to six months from hospital admission. The primary outcome was the occurrence of major amputation. For each classification system, the area under the receiver operating characteristic curve (AUROC) was calculated. Pairwise comparisons were performed to determine differences in prognostic performance, and the category or score with the highest discriminative value was identified for each classification.

**Results:** A total of 342 patients were included, with a mean age of 60 years; two-thirds were male. The cumulative incidence of major amputation at six months was 9.6%. The AUROC values were as follows: Saint Elian Classification, 0.90; Texas University, 0.81; Meggitt-Wagner, 0.80; and SINBAD, 0.74. The most predictive thresholds identified were a score of 18 for Saint Elian, stage 3D for the University of Texas, category 3 for Meggitt-Wagner, and a score of 5 for SINBAD.

**Conclusion:** The Saint Elian and University of Texas classifications demonstrated good prognostic accuracy for major amputation at six months, while the Meggitt-Wagner and SINBAD classifications showed moderate performance. Strengthening the training of multidisciplinary teams in referral centers is essential to ensure the effective application of these classification systems in clinical decision-making.

## Introduction

Diabetic foot is one of the most frequent and severe complications of diabetes mellitus. A lower limb amputation due to diabetic foot occurs every 20 seconds worldwide, making it the leading cause of non-traumatic limb loss [1]. Patients with diabetic foot ulcers have a 2.5-fold increased risk of death within five years compared to those with diabetes but without ulcers [2]. The direct costs associated with diabetic foot account for approximately one-third of total diabetes-related expenditures and are comparable to the investments made in oncological diseases [3]. Given its high burden, the study of its prevalence and prognosis is essential to improve clinical outcomes and optimize healthcare resource allocation [4].

In clinical practice, prediction rules based on signs, symptoms, medical history, and laboratory findings are widely used to assess disease prognosis, guide therapeutic decisions, and enhance communication between healthcare providers and patients [5,6]. In the case of diabetic foot, such tools are primarily applied to predict the likelihood of major amputation or the success of limb-salvage interventions [7]. Currently, more than 25 classification systems have been developed, each incorporating a different number of criteria [8]. Their performance varies depending on the available technology, the characteristics of the reference population, and the presence of multidisciplinary care teams [9].

In the local context, diabetic foot affects approximately 30% of patients attending healthcare facilities [10]. However, only 11.5% to 15% of these patients achieve adequate metabolic control, defined as optimal levels of blood glucose, arterial pressure, and lipid profile [11,12]. Furthermore, diabetic foot is the reason for hospital admission in one out of every five patients with diabetes in Peruvian hospitals [13]. Limitations in healthcare personnel training, infrastructure, and funding hinder the establishment of multidisciplinary teams dedicated to diabetic foot care, which contributes to suboptimal clinical outcomes.

Therefore, validating prognostic tools in settings different from those in which they were originally developed is essential to ensure their accuracy and applicability. Local contextual factors may influence the predictive performance of clinical scoring systems for major amputation. Considering the technology and healthcare conditions available locally, this study aimed to compare the discriminative capacity of four classification systems—Meggitt-Wagner (MW), University of Texas score (TU), SINBAD, and Saint Elian (SE)—to predict major amputation at six months of follow-up in patients managed at the Diabetic Foot Unit of a national hospital.

## Methods

### Study design and clinical scenario

A validation study was conducted using a retrospective cohort design based on data from the Diabetic Foot Unit of María Auxiliadora Hospital, covering the period from 2015 to 2019. This healthcare facility is classified as a Level III-1 referral hospital under the Peruvian Ministry of Health (MINSA) and is located in the southern area of Metropolitan Lima. It serves as a referral center for 13 surrounding districts, encompassing an estimated population of 2.5 million people, primarily insured by the public health system known as *Seguro Integral de Salud* (SIS). The hospital receives referrals from across southern Lima, including complex diabetic foot cases.

### Population, sample and sampling

The study included all patients with diabetic foot who were managed either on an outpatient basis or through hospitalization at María Auxiliadora Hospital between 2015 and 2019, with a minimum follow-up of six months from the date of admission. To be eligible, patients also needed to have complete clinical information within the first 48 hours of admission to allow for scoring according to the selected classification systems.

Exclusion criteria included foot lesions originating from peripheral venous insufficiency or pressure ulcers on the heel associated with prolonged immobilization or bedridden status.

All patients who met the eligibility criteria were included in the study. The statistical power of the resulting sample was calculated and is detailed in the analysis plan. A non-probabilistic, convenience sampling strategy was used, as the study relied on a pre-existing database of consecutively managed patients during the study period.

### Variables

#### Diabetic foot classifications

The components and categories of the four diabetic foot classifications used in this study were as follows:

#### Meggitt-Wagner Classification (MW)

This system is primarily based on ulcer depth, with partial consideration of ischemia. It includes six grades: grade 0, intact skin; grade 1, superficial ulcer; grade 2, ulcer reaching tendon or joint capsule; grade 3, deep ulcer with osteomyelitis or abscess; grade 4, localized gangrene (e.g., toes or forefoot); and grade 5, extensive gangrene involving the entire foot [14].

#### Texas University Classification (TU)

This system incorporates three dimensions: ulcer depth, infection, and ischemia. Depth is graded from 0 to 3: (0) pre-or post-ulcerative lesion with intact skin, (1) superficial wound not involving tendon, capsule or bone, (2) wound penetrating to tendon or capsule, and (3) wound penetrating to bone or joint. The presence of infection and/or ischemia is categorized as follows: (A) neither present, (B) infection, (C) ischemia, (D) both infection and ischemia. These combinations yield 16 possible grades with distinct prognostic implications for limb salvage and amputation [15].

#### SINBAD Classification

Developed by Ince et al. in 2008 as a modification of the (S[AD])SAD system, it is an acronym that evaluates six parameters: *Site* (forefoot vs. midfoot/hindfoot), *Ischemia* (yes/no), *Neuropathy* (yes/no), *Bacterial infection* (yes/no), *Area* (<1 cm² or ≥1 cm²), and *Depth* (superficial or involving muscle/bone). This simplified score is intended for broad applicability in clinical and resource-limited settings [16].

#### Saint Elian Classification (SE)

This system is composed of ten parameters organized into three domains.

- *Anatomical factors*: location (phalanges, metatarsal, or tarsal), topography (dorsal/plantar, lateral/medial, or involving two or more areas), and number of affected regions (one, two, or the entire foot).
- *Aggravating factors*: ischemia (none, mild, moderate, or severe), infection (none, mild, moderate, or severe), edema (none, perilesional, limited to the affected leg, or bilateral), and neuropathy (none, diminished sensation, complete loss of protective sensation, or presence of Charcot arthropathy).
- *Contributing factors*: depth (superficial, below dermis, or full-thickness), ulcer area (<10 cm², 10–40 cm², or >40 cm²), and healing phase (epithelialization, granulation, or inflammatory phase).

A cumulative score is generated, classifying severity as mild (<10 points), moderate (11– 20 points), or severe (21–30 points) [17].

### Major amputation

Major amputation was defined as any amputation performed above the ankle joint, including infracondylar and supracondylar levels.

### Other variables

The following covariates were considered in the analysis:

- **Sociodemographic variables:** Age in years (<60; ≥60), sex (male/female), and educational attainment (primary or less; secondary; higher education).
- **Anthropometric and clinical data:** Body mass index (BMI) in kg/m² (<25; 25–29; ≥30), diabetes duration in years (<10; ≥10), and type of diabetes treatment (diet only; oral antidiabetic drugs; insulin).
- **Medical history:** Presence or absence of hypertension, coronary artery disease, stroke, and history of prior major amputation.
- **Ulcer characteristics:** Type (new vs. recurrent), size (measured 48 hours after admission using the ellipse formula, categorized as <10 cm²; 10–39 cm²; ≥40 cm²), and depth (based on the IWGDF PEDIS classification). Ulcer location was classified as forefoot (yes/no).
- **Ischemia:** Defined by arterial Doppler findings (presence of biphasic, monophasic, or absent flow in the anterior tibial, posterior tibial, or popliteal arteries).
- **Laboratory data:** Glycated hemoglobin (HbA1c <7%; ≥7%), hemoglobin (<8 g/dL; ≥8 g/dL), LDL cholesterol (<100 mg/dL; ≥100 mg/dL), and serum albumin (<2.5 g/dL; ≥2.5 g/dL).

### Procedures

#### Preparation

Permission to access the diabetic foot database was obtained from the Endocrinology Service of María Auxiliadora Hospital. The dataset was provided in a fully anonymized format, with all personally identifiable information removed to ensure patient confidentiality. Subsequently, the records were filtered and depurated according to the predefined eligibility criteria for this study.

### Diagnostic and therapeutic algorithm at the María Auxiliadora hospital

At the hospital, all patients with diabetes mellitus presenting with lower limb ulcers, infections, or ischemia—whether as outpatients or via the emergency department—are evaluated by the Diabetic Foot Unit to determine the need for outpatient care or hospitalization. Upon admission, the Unit systematically registers patient data and documents each debridement and wound care session.

Management protocols follow the guidelines established by the International Working Group on the Diabetic Foot (IWGDF) [18]. Initial antibiotic therapy typically involves broad-spectrum agents, subsequently tailored based on antibiogram results obtained from tissue biopsies collected during the first surgical debridement, performed by Unit personnel.

Patients with more severe conditions undergo daily wound care from Monday to Saturday during the acute phase. This frequency is reduced to three times per week once clinical stabilization is achieved, continuing until hospital discharge. Both outpatients and those discharged from inpatient care are followed up weekly in the Unit until complete epithelialization is achieved.

Noninvasive vascular assessments are performed using arterial Doppler ultrasound. Ischemia is diagnosed based on the presence of monophasic or biphasic waveforms in the anterior tibial, posterior tibial, or popliteal arteries of the affected limb.

All patients receive intensive insulin therapy to maintain glycemic targets between 140 and 180 mg/dL, along with high-protein and hypoglycemic diets. Anemia is actively managed, including blood transfusions when clinically indicated.

### Statistic analysis

The authors accessed and processed the dataset between December 1, 2022 and Augut 31, 2023. All analyses were conducted using R software, version 2025.05.0+496. Statistical power was calculated for each of the six comparisons performed, assuming a two-sided alpha level of 5%, with 39 cases and 303 controls. Power estimations based on AUROC values are presented in Supplementary Table 2.

Descriptive statistics were reported using absolute and relative frequencies for categorical variables, stratified by the presence or absence of major amputation. Differences between groups were assessed using the chi-square test for nominal variables and the Wilcoxon rank-sum test for ordinal or non-normally distributed variables. Additionally, the median and interquartile range (IQR) of length of hospital stay were calculated.

The cumulative six-month incidence of major amputation was determined according to MW, TU, SINBAD, and SE. For each, relative risk (RR) and 95% confidence intervals (CI) for major amputation were calculated across all categories, using the lowest theoretical risk category as reference. Categories with zero observations were collapsed with adjacent ones as appropriate. For TU, each component (depth, infection, and ischemia) was evaluated separately due to data concentration in stage 3D versus other categories.

AUROC was calculated for each classification to assess overall discriminatory ability and compared using the DeLong test for correlated ROC curves. Sensitivity, specificity, positive predictive value (PPV), and negative predictive value (NPV) were estimated for each category to predict major amputation. Optimal cut-off points were identified using the Youden Index. A two-sided p-value < 0.05 was considered statistically significant.

### Ethical aspects

This study was approved by the Endocrinology Department of María Auxiliadora Hospital and the Institutional Ethics and Research Committee of the Universidad Científica del Sur (Approval Certificate No. 386-CIEI-CIENTÍFICA-2021). The analyzed database contained no personally identifiable information, as all data were encrypted. Only physicians from the hospital’s endocrinology service had access to the identification codes linking patient data.

## Results

### General characteristics

The average age of the population was 60 years, with two out of three being men. Hypertension was present in 38%, 3.9% had a prior stroke, and 7% had a previous major amputation. Diabetes duration was ≥ 10 years in 62%, 41% received insulin treatment before infection, 80% had HbA1c ≥ 7%, 12% had albumin < 2.5 g/dL, and 75% had hemoglobin < 8 g/dL. Regarding foot characteristics, 77% had an ulcer extension greater than 40 cm², 97% presented a new ulcer, 59% had ischemia, and 46% had severe infection. The major amputation rate was 11% (Table 1).

**Table 1.**
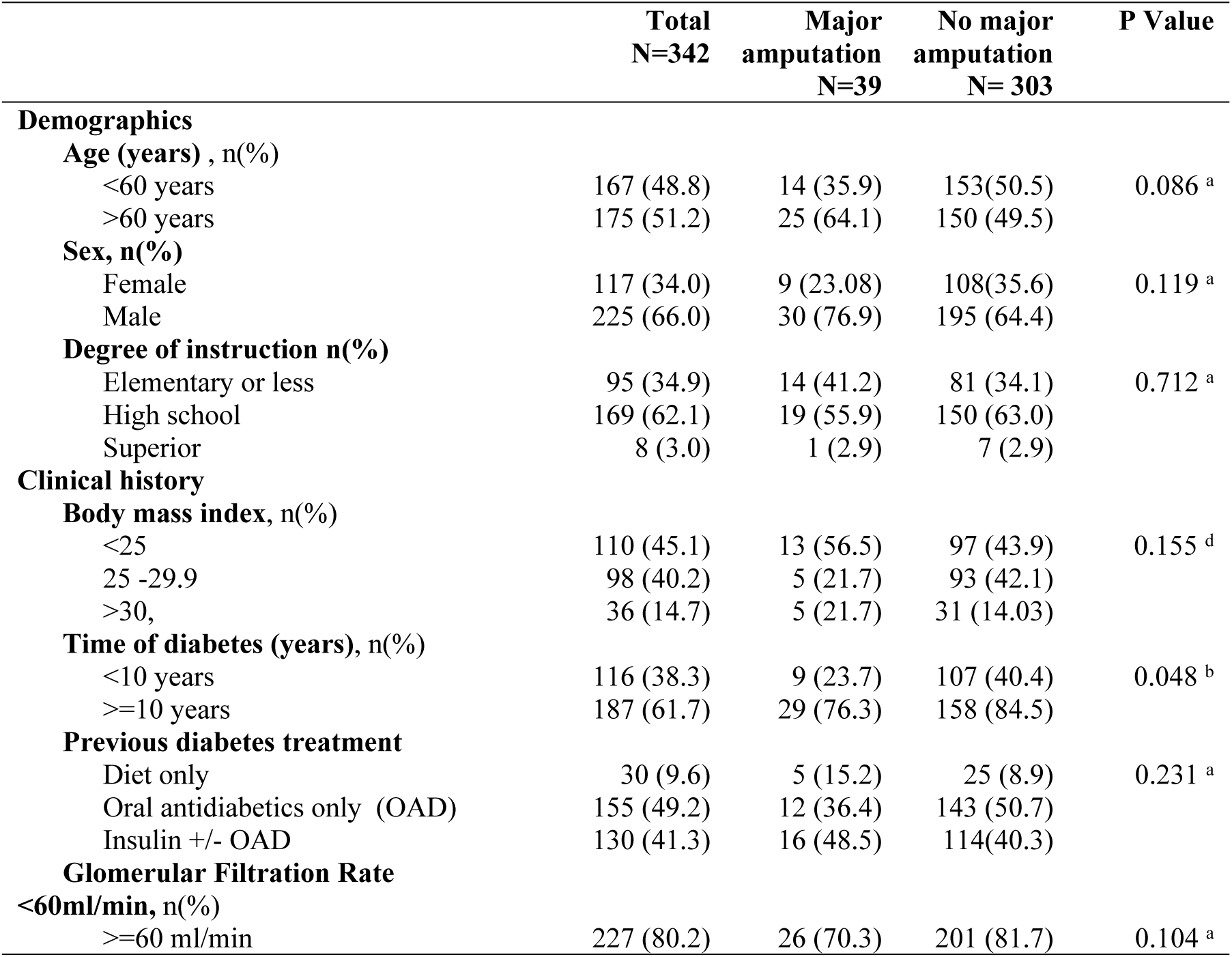
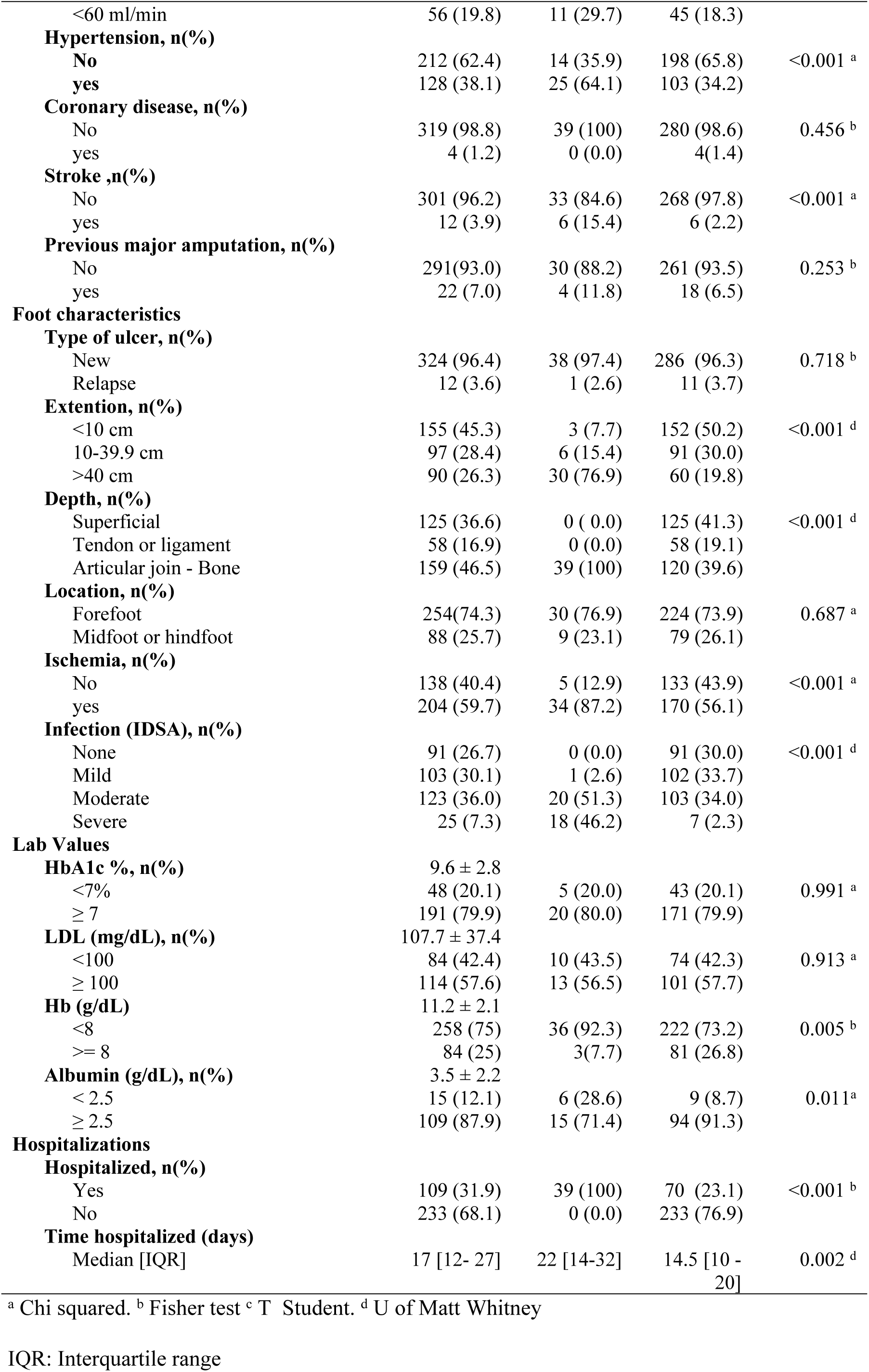
Incidence of major amputation according to clinical epidemiological characteristics of patients with diabetic foot at the Maria Auxiliadora Hospital.

### Differences between amputees and non-amputees

Patients with major amputation had a longer duration of illness and higher prevalence of comorbidities such as hypertension and prior stroke. Age, education level, and body mass index were similar between groups.

Regarding diabetic foot features, major amputation was associated with greater ulcer depth, severe infection, ischemia, and larger ulcer extension, but not with ulcer location or recurrence. Laboratory variables showed an association between albumin < 2.5 g/dL and major amputation, whereas HbA1c, LDL cholesterol, and hemoglobin showed no significant association (Table 1).

### Risk of major amputation

MW grades 4 or 5 increased the risk of major amputation 14.7 times (RR 15.7; 95% CI 4.9 to 50.2; p < 0.001) compared to the combined categories 1–3. TU stage 3D increased the risk 13.3 times (RR 14.3; 95% CI 5.7 to 35.7; p < 0.001) compared to the rest of the combined stages. When analyzing the three individual components of TU, only ischemia had sufficient cases to calculate risk (RR 4.59; 95% CI 1.84 to 11.48). A SINBAD score of 5 increased the risk 6.88 times (RR 7.88; 95% CI 3.11 to 19.9) compared to scores less than 4. Finally, an SE score greater than 20 increased the risk 31.2 times (95% CI 10.0 to 103.6) compared to scores less than 16 (Table 2).

**Table 2.** Incidence of major amputation according to categories of the 4 diabetic foot classifications.

|  | Major<br>amputation | No major<br>amputation | Relative<br>risk | CI 95% | P Value |
| --- | --- | --- | --- | --- | --- |
| **Meggitt -Wagner scale, n(%)** |  |  |  |  |  |
| 1-3 | 3 (1.6) | 191 (98.5) | 1.0 |  |  |
| 4-5 | 36 (24.3) | 112 (75.7) | 15.7 | 4.91 – 50.2 | 0.001 |
| **Texas University -Depth, n(%)** |  |  |  |  |  |
| Superficial | 0 (0.0) | 125 (100.0) | NC |  |  |
| Tendon or ligament | 0 (0.0) | 58 (100.0) |  |  |  |
| Bone | 39 (24.5) | 120 (75.5) |  |  |  |
| **Texas University – infection, n(%)** |  |  |  |  |  |
| No | 0 (0.0) | 91 (100.0) | NC |  |  |
| Yes | 39 (15.5) | 212 (84.5) |  |  |  |
| **Texas University – ischemia, n(%)** |  |  |  |  |  |
| No | 5 (3.6) | 133 (96.4) | 1.00 |  |  |
| Yes | 34 (16.7) | 170 (83.3) | 4.59 | 1.84- 11.48 | 0.001 |
| **Texas University – 3D, n(%)** |  |  |  |  |  |
| 1a – 3c | 5 (2.2) | 227 (97.8) | 1.0 |  |  |
| 3d | 34 (30.9) | 76 (69.1) | 14.3 | 5.75 – 35.7 | <0.001 |
| **Sinbad score, n(%)** |  |  |  |  |  |
| 1-4 | 5 (2.8) | 177 (97.3) | 1.00 |  |  |
| 5 | 26 (21.7) | 94 (78.3) | 7.88 | 3.11 – 19.9 | <0.001 |
| 6 | 8 (20.0) | 32 (80.0) | 7.28 | 2.50 – 21.1 | <0.001 |
| **Saint Elia score, n(%)** |  |  |  |  |  |
| <16 | 3 ( 1.5) | 200 (98.5) | 1.0 |  |  |
| 16 -20 | 16 (16.5) | 81 (83.5) | 11.1 | 3.32 – 37.4 | <0.001 |
| >20 | 20 (47.6) | 22 (52.4) | 32.2 | 10.0 – 103.6 | <0.011 |

NC:Not calculated

### Prognostic capacity

Seven ROC curves corresponding to four classification systems—SE, MW, TU (3D stage, depth, ischemia, infection), and SINBAD—were analyzed. **(Figure 1)**

**Figure 1.**
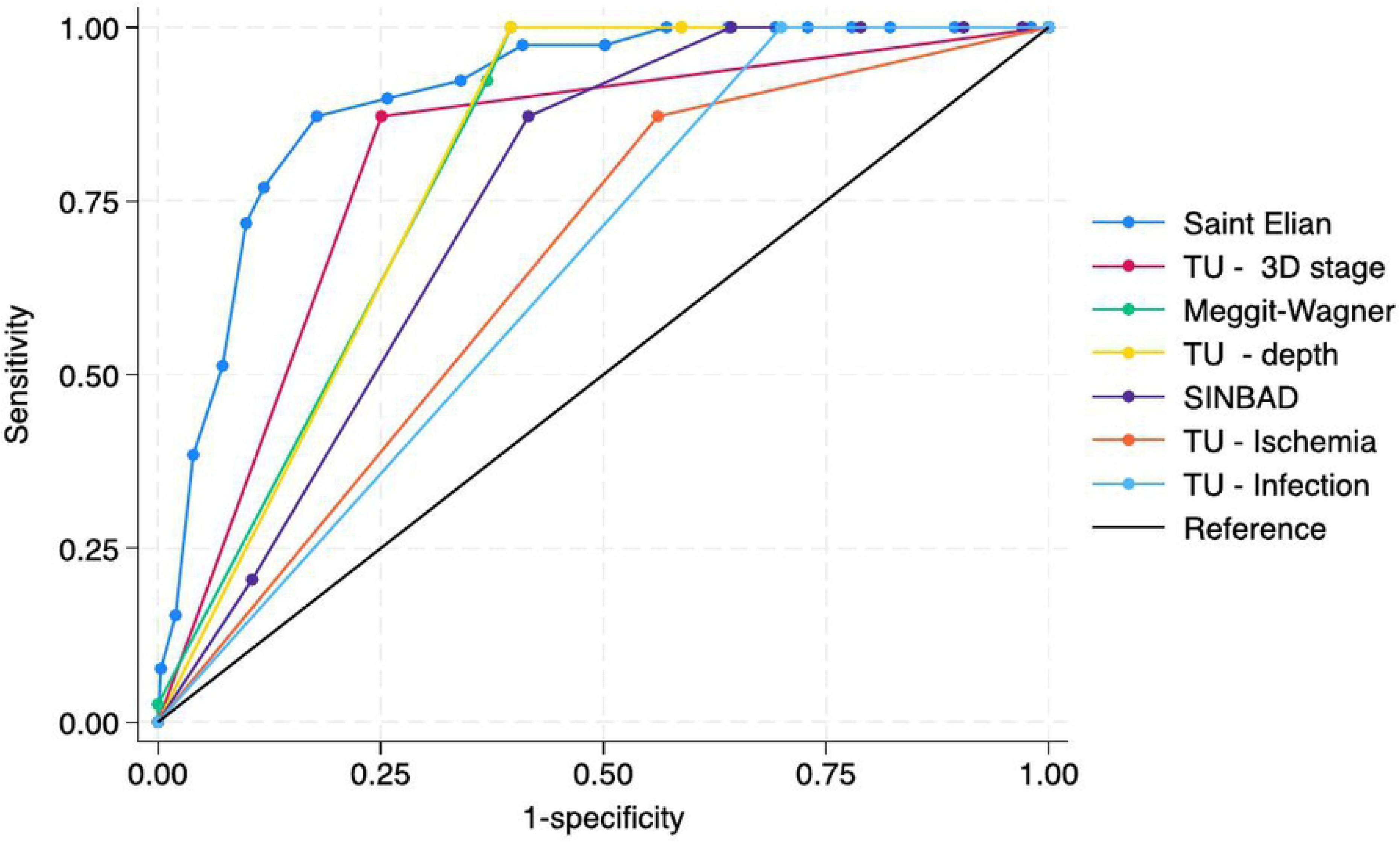
Comparison of the Areas under the ROC curve of the 4 diabetic foot classifications for major amputation of patients. Areas under the curve. Saint Elian 0.900 (95%CI 0.858 – 0.942); TU-3d stage, 0.811 (95%CI 0.752-0.869); Meggitt Wagner 0.805 (95%CI 0.770 – 0.839); TU depth 0.802 (0.774 – 0.29); SINBAD 0.747 (95%CI 0.693 – 0.802); TU ischemia 0.655 (95%CI 0.595 – 0.715); TU infection 0.650 (95% CI 0.624 – 0.676).

The SE scale demonstrated the best accuracy compared to the others, with an AUROC of 0.900. Followed by TU 3D stage (AUROC = 0.811) and MW (AUROC = 0.805), both showing similar performance. Lastly, SINBAD had the lowest discriminative ability, with an AUROC of 0.747. Regarding the individual components of UT, depth showed an accuracy similar to MW (AUROC = 0.802). Meanwhile, the ischemia and infection components exhibited lower performances, with AUROCs of 0.655 and 0.650, respectively, being the lowest among all **(Table 3).**

**Table 3.**
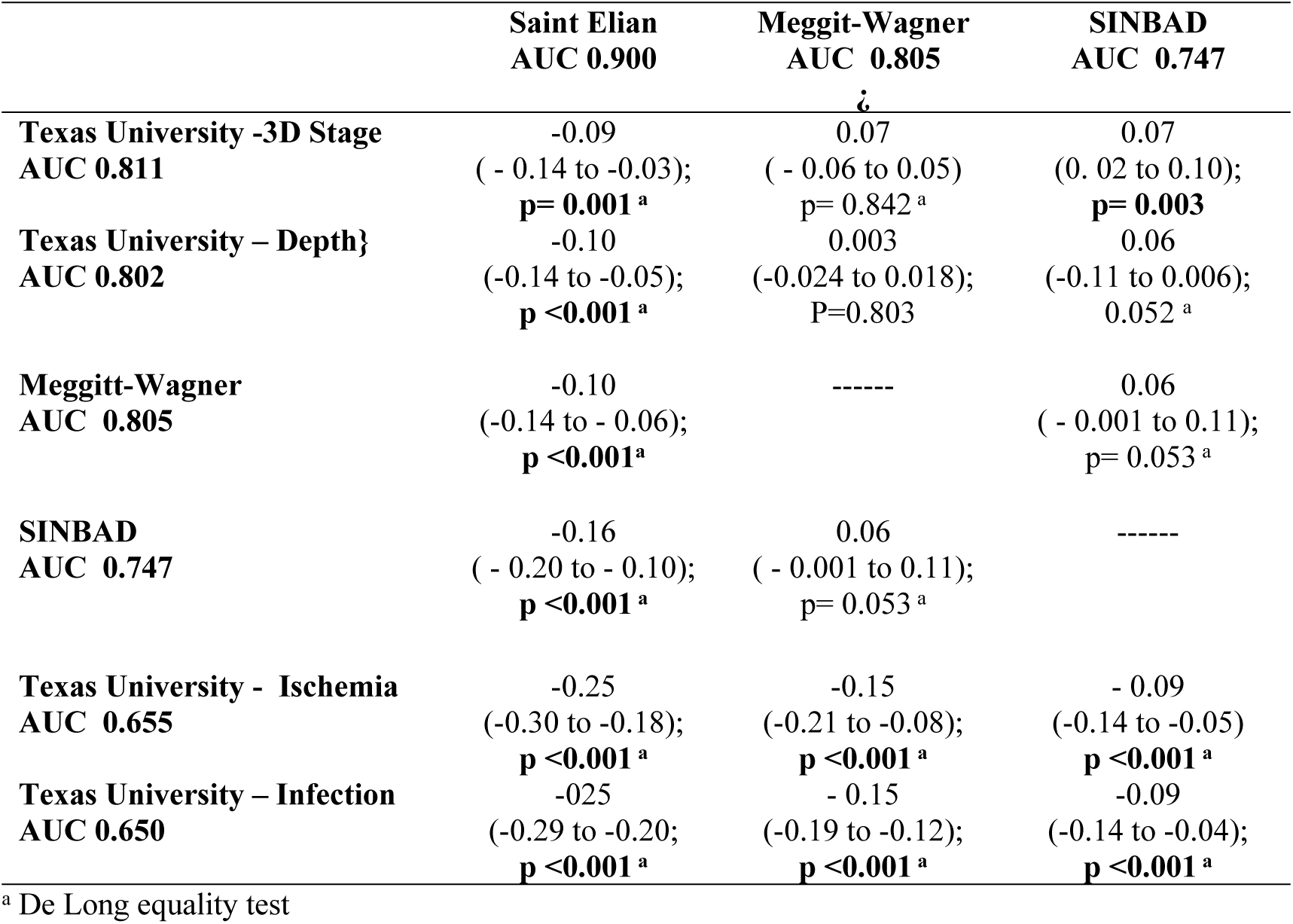
Differences in areas under the ROC curve of the prognostic capacity for major amputation between diabetic foot scales.

The performance of the classifications was informed thanks to the Youden index of each one. A Wagner score greater than or equal to 3 had an index of 52.7%, a Texas 3D stadium had an index of 62.1%, a SINBAD score greater than or equal to 5 had an index of 45.6%, and a Saint Elian score greater than or equal to 18 had an index of 69.4 **(Table 4)**

**Table 4.** Prognostic discriminative capacity of cut off points of the diabetic foot classifications for major amputation in patients at the Maria Auxiladora hospital.

|  | Sensitivity (95% CI) | Specificity (95% CI) | PPV (95% CI) | NPV (95% CI) | LR [+] (95% CI) | LR [-] (95% CI) | Youden index |
| --- | --- | --- | --- | --- | --- | --- | --- |
| **Meggitt-Wagner** |  |  |  |  |  |  |  |
| ≥ 3 | **100.0 (91-100)** | **60.4 (54.6-65.9)** | **24.5 (18.1-32)** | **100.0 (98-100)** | **2.52 (2.2-2.9)** | **0.00** | **60.4** |
| ≥ 4 | 92.3 (79.1-98.4) | 63.0 (57.3-68.5) | 24.3 (17.7-32.1) | 98.5 (95.5-99.7) | 2.49<br>(2.1-2.97) | 0.12<br>(0.041-0.363) | 55.3 |
| **TU Depth** |  |  |  |  |  |  |  |
| ≥ 2 | 100 (91-100) | 41.3 (35.7-47) | 17.9 (13.1-23.7) | 100.0 (97.1-100) | 1.70 (1.55-1.87) | 0.00 | 41.3 |
| ≥ 3 | **100 (91-100)** | **60.4 (54.6-65.9)** | **24.5 (18.1-32)** | **100.0 (98-100)** | **2.52 (2.2-2.9)** | **0.00** | **60.4** |
| **TU Ischemia** |  |  |  |  |  |  |  |
| **Yes** | 87.2 (72.6-95.7) | 43.9 (38.2-49.7) | 16.6 (11.8-22.5) | 96.4 (91.7-98.8) | 1.55 (1.33-1.82) | 0.29 (0.12-0.66) | 31.1 |
| **TU infection** |  |  |  |  |  |  |  |
| **Yes** | 100.0 (91-100) | 30.0 (24.9-35.5) | 15.5 (11.3-20.6) | 100.0 (96-100) | 1.43 (1.33-1.54) | 0.00 | 30.0 |
| **TU 3D** |  |  |  |  |  |  |  |
| **3D** | **87.2 (72.6-95.7)** | **74.9 (69.6-79.7)** | **30.9 (22.4-40.4)** | **97.8 (95-99.3)** | **3.47 (2.76-4.37)** | **0.17 (0.07-0.38)** | **62.1** |
| ≥ 4 | 100.0 (91-100) | 35.6 (30.2-41.3) | 16.7 (12.1-22.1) | 100.0 (96.6-100) | 1.55 (1.43-1.69) | 0.00 | 35.6 |
| ≥ 5 | **87.2 (72.6-95.7)** | **58.4 (52.6-64)** | **21.3 (15.2-28.4)** | **97.3 (93.7-99.1)** | **2.09 (1.75-2.51)** | **0.21 (0.09-0.5)** | **45.6** |
| **Saint Elian** |  |  |  |  |  |  |  |
| ≥ 10 | 100.0 (91-100) | 22.1 (17.6-27.2) | 15 (10.3-18.9) | 100.0 (95.6-100) | 1.37 (1.28-1.47) | 0.00 | 27.1 |
| ≥ 15 | 97.4 (86.5-99.9) | 59.1 (53.3-64.7) | 23.5 (17.2-30.7) | 99.4 (96.8-100) | 2.38 (2.06-2.75) | 0.04 (0.06-0.30) | 56.5 |
| ≥ 17 | 89.7 (75.8-97.1) | 74.3 (68.9-79.1) | 31.0 (22.6-40.4) | 98.3 (95.6-99.5) | 3.48 (2.8-4.3) | 0.13 (0.05-0.35) | 64.0 |
| ≥ 18 | **87.2 (72.6-95.7)** | **82.2 (77.4-86.4)** | **38.6 (28.4-49.6)** | **98.0 (95.5-99.4)** | **4.89 (3.73-6.41)** | **0.15 (0.68-0.35)** | **69.4** |
| ≥ 20 | 71.8 (55.1-85) | 90.1 (86.2-93.2) | 48.3 (35-61.8) | 96.1 (93.2-98.1) | 7.25 (4.9-10.7) | 0.31 (0.18-0.51) | 61.9 |

PV: Positive predictive value. NPV: negative predictive value. LR(+). Positive likelihood ratio LR (-) Negative likelihood ratio

## Discussion

### Main findings

Knowing the performance of prognostic tools is essential in contexts with limited resources. Our study found that at six months of follow-up, the prognostic capacity of the Saint Elian and Texas classifications was good, while that of the Wagner and SINBAD classifications was moderate. The most discriminative cut-off points were a score of 18 for Saint Elian, stage 3D for Texas, category 4 for Wagner, and a score of 5 for SINBAD.

### Comparison with previous studies

#### Saint Elian classification

At the Latin American level, our results are consistent with an Argentine study that followed patients with new-onset diabetic foot ulcers for five months. This study reported an AUC of 0.893 for major amputation using the Saint Elian classification with a cut-off point greater than 18 [19]. Similarly, a Chinese study found that a Saint Elian score greater than 17 reduced the probability of cure by 24%, although it did not report statistics for major amputation alone due to a small sample size [20]. The Saint Elian system, composed of ten variables, is theoretically better suited to predict major amputation; however, the inclusion of multiple variables increases interobserver variability and may lead to inconsistent results. This highlights the importance of training and standardized validation procedures.

### Texas University Classification

Regarding the UT classification, a Philippine study reported an AUC of 0.785 for grade and 0.575 for stage [21]. In contrast, in our analysis, the University of Texas classification showed the second-best performance (AUC = 0.81) for major amputations. This difference may be explained by our dichotomous analysis, focused specifically on the presence of stage 3D, due to its high prevalence in our hospital population. The Texas system does not yield a single score but instead assigns patients into one of 16 categories based on four grades and four stages [22]. In practice, the grade assesses lesion depth, while the stage identifies infection or ischemia. An expanded version, the WIfI classification, was introduced nine years ago and also incorporates infection and ischemia categorization into a single outcome. However, we were unable to evaluate this system due to limitations in vascular assessment [23].

### Wagner Classification

The Wagner classification demonstrated intermediate prognostic performance. It primarily assesses ulcer depth and includes only a partial assessment of peripheral arterial disease. In cases with necrosis, it cannot distinguish whether the cause is infection or ischemia. As one of the earliest and most widely used classification systems, Wagner’s utility in research is limited by its lack of detail regarding infection severity or ischemic status [24]. Nonetheless, in our study population, which included many patients with deep lesions and necrosis, this system had the third-best performance.

### SINBAD Classification

The SINBAD classification, composed of six dichotomized variables, had the lowest performance (AUC = 0.74). While this system is endorsed by the International Working Group on the Diabetic Foot (IWGDF) for communication among healthcare providers and outcome comparisons across institutions [25], its binary structure and fixed cut-offs may limit its discriminative power in high-risk populations such as hospitalized patients.

### Theoretical and practical implications

After development of MW classification, other systems included depth, infection, and ischemia categories to improve prognostic accuracy. Authors have no limits to expand variables or modify category definitions [26]. Although a greater number of variables may improve performance in theory, it also increases the potential for interobserver variability. Therefore, proper training and standardization of classification methods are critical to reduce heterogeneity and ensure reliable prognostic evaluation [27].

A recent systematic review aimed at evaluating prognostic tools for amputation could not provide pooled outcomes by classification systems due to heterogeneity in included studies and inconsistencies in the measurement of the same tools. The reviewed studies lacked sufficient methodological rigor, underscoring the need for future research to adopt standardized definitions and training protocols to ensure the external validity of outcomes in clinical practice [28].

### Public health importance

The classifications can have different uses in research, auditing, clinical monitoring and prognosis. The estimates found in this study may change depending on the population evaluated, the resources of the health system and the existence of a diabetic foot management program. At the primary level, the Peruvian diabetic foot guidelines consider only the Wagner classification for reference purposes [29]. World guidelines suggest using SINBAD because it is simple and easy to use, and could be an alternative to improve the communication and statistics [18].

We do not have diabetic foot national guidelines for hospital setting. However, worldwide there is a great variety in which the choice will depend on the type of patient, the available resources and, above all, having a multidisciplinary team for the diagnosis and management of diabetic foot, which even with limited resources and minimal equipment, can achieve significant changes in the amputation rate [30].

### Strengths and limitations

Within the limitations, the performance of the classifications cannot be extrapolated to the population of other hospital centers in Peru, so validations considering multiple centers are necessary. Vascular evaluation was only performed by plethysmography of the pedis and tibial arterial waves. This form of evaluation is contemplated in the guidelines for peripheral arterial disease and the diabetic foot classifications, only the Saint Elian classification considers it[17]. Its extensive use requires validation studies in resource-limited contexts like ours.

Among the strengths, a minimum follow-up of 6 months was carried out to evaluate the occurrence of the outcome due to persistence of the initial injury. Despite being retrospective, all patients who were hospitalized were considered without exception and the doctors of the endocrinology service carried out an active search for the outcomes, even if the patient did not continue with the post-acute event evaluations.

### Conclusion

In patients with infected diabetic foot from a Peruvian hospital, the six-month prognosis for major amputation of the Texas and Saint Elian classifications was good and that of the Wagner and SINBAD classifications was moderate. It is a priority to reinforce the training of multidisciplinary teams in reference centers to efficiently use the results of any of these classifications. Each facility must determine its own prognostic capacity based on available resources and determine the most useful classification for the type of population it serves.

## List of Supplementary tables

**S1 Table.** Sample power of comparison between AUROC curves

**S2 Table.** Incidence of major amputation according to detailed diabetic foot classifications

**S3 table.** Prognostic discriminative capacity of cut off points of the diabetic foot classifications for major amputation in patients at the Maria Auxiladora hospital

## Data Availability

All relevant data are within the manuscript and its Supporting Information files.

## Abbreviations

SE: Saint Elian
MW: Meggitt-Wagner
TU: University of Texas score
AUROC: Area under the receiver operating characteristic curve
MINSA: Peruvian Ministry of Health
SIS: Seguro Integral de Salud
IWGDF: International Working Group on the Diabetic Foot

## Bibliographical references

1. Asociación Latinoamericana de Diabetes. Guías ALAD sobre el Diagnóstico, Control y Tratamiento de la Diabetes Mellitus Tipo 2 con Medicina Basada en Evidencia. 2019. Available from: https://www.revistaalad.com/guias/5600AX191_guias_alad_2019.pdf

2. Walsh JW, Hoffstad OJ, Sullivan MO, Margolis DJ. Association of diabetic foot ulcer and death in a population-based cohort from the United Kingdom. Diabet Med. 2016; 33(11): 1493–8. doi:10.1111/dme.13054

3. Armstrong DG, Swerdlow MA, Armstrong AA, Conte MS, Padula WV, Bus SA. Five year mortality and direct costs of care for people with diabetic foot complications are comparable to cancer. J Foot Ankle Res. 2020; 13(1): 2–5. doi:10.1186/s13047-020-00383-2

4. Shaw JE, Sicree RA, Zimmet PZ. Global estimates of the prevalence of diabetes for 2010 and 2030. Diabetes Res Clin Pract. 2010; 87: 4–14. doi:10.1016/j.diabres.2009.10.007

5. Wasson JH, Sox HC, Neff RK, Goldman L. Clinical prediction rules, Applications and methodological standards. N Engl J Med. 1985; 313(13): 793–9. doi:10.1056/NEJM198509263131306

6. Jeffcoate WJ, Macfarlane RM, Fletcher EM. The Description and Classification of Diabetic Foot Lesions. Diabet Med. 1993; 10(7): 676–9. doi:10.1111/j.1464-5491.1993.tb00144.x

7. Armstrong DG, Peters EJ. Classification of wounds of the diabetic foot. Curr Diab Rep. 2001; 1(3): 233–8. doi:10.1007/s11892-001-0039-1

8. Games F. Classification of diabetic foot ulcers. Diabetes Metab Res Rev. 2016; 32: 186–94. doi:10.1002/dmrr.2746

9. Ramos W, López T, Revilla L, More L, Huamaní M, Pozo M, et al. Resultados de la vigilancia epidemiológica de diabetes mellitus en hospitales notificantes del Perú, 2012. Rev Peru Med Exp Salud Publica. 2014; 31(1): 9–15. PMID:24718521

10. Azañedo D, Bendezú-Quispe G, Lazo-Porras M, Cárdenas-Montero D, Beltrán-Ale G, Thomas NJ, et al. Calidad de control metabólico en pacientes ambulatorios con diabetes tipo 2 atendidos en una clínica privada. Acta Med Peru. 2017; 34(2): 106–19. ISSN:1728-5917

11. Huayanay-Espinoza IE, Guerra-Castañon F, Lazo-Porras M, Castaneda-Guarderas A, Thomas NJ, Garcia-Guarniz AL, et al. Metabolic control in patients with type 2 diabetes mellitus in a public hospital in Peru: A cross-sectional study in a low-middle income country. PeerJ. 2016; 2016(10). doi:10.7717/peerj.2577

12. Yovera-Aldana M, Sáenz-Bustamante S, Quispe-Landeo Y, Agüero-Zamora R, Salcedo J, Sarria C, et al. Nationwide prevalence and clinical characteristics of inpatient diabetic foot complications: A Peruvian multicenter study. Prim Care Diabetes. 2021 Jun 1; 15(3): 480–7. doi:10.1016/j.pcd.2021.02.009

13. Meggit B. Surgical management of the diabetic foot. Br J Hosp Med. 1976; 16: 227–332. *Clin Calcium*. 2003 Sep; 13(9): 1179–84. PMID:15775200

14. Lavery LA, Armstrong DG, Harkless LB. Classification of diabetic foot wounds. J Foot Ankle Surg. 1996; 35(6): 528–31. doi:10.1016/s1067-2516(96)80125-6

15. Ince P, Abbas ZG, Lutale JK, Basit A, Ali SM, Chohan F, et al. Use of the SINBAD Classification System and Score in Comparing Outcome of Foot Ulcer Management on Three Continents. Diabetes Care. 2008; 31(5): 964–7. doi:10.2337/dc07-2367

16. Martínez-De Jesús FR. A checklist system to score healing progress of diabetic foot ulcers. Int J Low Extrem Wounds. 2010; 9(2): 74–83. doi:10.1177/1534734610371594

17. Schaper NC, van Netten JJ, Apelqvist J, Bus SA, Hinchliffe RJ, Lipsky BA, et al. Practical Guidelines on the prevention and management of diabetic foot disease. Diabetes Metab Res Rev. 2020; 36(S1): 1–10. doi:10.1002/dmrr.3266

18. Carro GV, Saurral R, Carlucci E, Gette F, Llanos M de los Á, Amato PS. A Comparison Between Diabetic Foot Classifications WIfI, Saint Elian, and Texas: Description of Wounds and Clinical Outcomes. Int J Low Extrem Wounds. 2022 Jun 1; 21(2): 120–30. doi:10.1177/1534734620930171

19. Huang Y, Xie T, Cao Y, Wu M, Yu L, Lu S, et al. Comparison of two classification systems in predicting the outcome of diabetic foot ulcers: The Wagner grade and the Saint Elian Wound score systems. Wound Repair Regen. 2015 May 1; 23(3): 379–85. doi:10.1111/wrr.12289

20. Vera-Cruz PN, Palmes PP, Tonogan LJM, Troncillo AH. Comparison of WIFi, University of Texas and Wagner Classification Systems as Major Amputation Predictors for Admitted Diabetic Foot Patients: A Prospective Cohort Study. Malays Orthop J. 2020; 14(3): 114–23. doi:10.5704/MOJ.2011.018

21. Armstrong DG, Lavery LA, Harkless LB. Validation of a Diabetic Wound Classification System: The contribution of depth, infection, and ischemia to risk of amputation. Diabetes Care. 1998; 21(5): 855–9. doi:10.2337/diacare.21.5.855

22. Mills JL, Conte MS, Armstrong DG, Pomposelli FB, Schanzer A, Sidawy AN, et al. The society for vascular surgery lower extremity threatened limb classification system: Risk stratification based on Wound, Ischemia, and foot Infection (WIfI). J Vasc Surg. 2014; 59(1): 220–34.e2. doi:10.1016/j.jvs.2013.08.003

23. Gonzales De La Torre H, Mosquera Fernández A, Quintana Lorenzo L, Perdomo Perez E, Montesdeoca MDPQ. Classifications of injuries on diabetic foot: A non-solved problem. Gerokomos. 2012; 23(2): 75–87. doi:10.4321/S1134-928X2012000200006

24. Monteiro-Soares M, Russell D, Boyko EJ, Jeffcoate W, Mills JL, Morbach S, et al. Guidelines on the classification of diabetic foot ulcers (IWGDF 2019). Diabetes Metab Res Rev. 2020 Mar 1; 36(S1). doi:10.1002/dmrr.3273

25. Monteiro-Soares M, Boyko EJ, Jeffcoate W, Mills JL, Russell D, Morbach S, et al. Diabetic foot ulcer classifications: A critical review. Diabetes Metab Res Rev. 2020; 36(S1): 1–16. doi:10.1002/dmrr.3272

26. Monteiro-Soares M, Martins-Mendes D, Vaz-Carneiro A, Dinis-Ribeiro M. Lower-limb amputation following foot ulcers in patients with diabetes: Classification systems, external validation and comparative analysis. Diabetes Metab Res Rev. 2015 Jul 1; 31(5): 515–29. doi:10.1002/dmrr.2634

27. Monteiro-Soares M, Martins-Mendes D, Vaz-Carneiro A, Sampaio S, Dinis-Ribeiro M. Classification systems for lower extremity amputation prediction in subjects with active diabetic foot ulcer: A systematic review and meta-analysis. Diabetes Metab Res Rev. 2014; 30(7): 610–22. doi:10.1002/dmrr.2535

28. Ministerio de Salud del Perú. Guía técnica: Guía de práctica clínica para el diagnóstico, tratamiento y control del pie diabético. Dirección General de Intervenciones Estratégicas en Salud Pública. 2017; 1–29 [cited 2025 Jun 13]. Available from: https://bvs.minsa.gob.pe/local/MINSA/3971.pdf

29. Karthikesalingam A, Holt PJE, Moxey P, Jones KG, Thompson MM, Hinchliffe RJ. A systematic review of scoring systems for diabetic foot ulcers. Diabet Med. 2010; 27(5): 544–9. doi:10.1111/j.1464-5491.2010.02989.x

